# Pre-existing conditions are associated with COVID patients’ hospitalization, despite confirmed clearance of SARS-CoV-2 virus

**DOI:** 10.1101/2020.10.28.20221655

**Authors:** Colin Pawlowski, AJ Venkatakrishnan, Eshwan Ramudu, Christian Kirkup, Arjun Puranik, Nikhil Kayal, Gabriela Berner, Akash Anand, Rakesh Barve, John C. O’Horo, Andrew D. Badley, Venky Soundararajan

## Abstract

The current diagnostic gold-standard for SARS-CoV-2 clearance from infected patients is two consecutive negative PCR test results. However, there are anecdotal reports of hospitalization from protracted COVID complications despite such confirmed viral clearance, presenting a clinical conundrum. We conducted a retrospective analysis of 266 COVID patients to compare those that were admitted/re-admitted post-viral clearance (*hospitalized post-clearance cohort*, n=93) with those that were hospitalized pre-clearance but were not re-admitted post-viral clearance (*non-hospitalized post-clearance cohort*, n=173). In order to differentiate these two cohorts, we used neural network models for the augmented curation of comorbidities and complications with positive sentiment in the EHR physician notes. In the year preceding COVID onset, acute kidney injury (n=15 (16.1%), p-value: 0.03), anemia (n=20 (21.5%), p-value: 0.02), and cardiac arrhythmia (n=21 (22.6%), p-value: 0.05) were significantly enriched in the physician notes of the *hospitalized post-clearance cohort*. This study highlights that these specific pre-existing conditions are associated with amplified hospitalization risk in COVID patients, despite their successful SARS-CoV-2 viral clearance. Our finding that pre-COVID anemia amplifies risk of post-COVID hospitalization is particularly concerning given the high prevalence and endemic nature of anemia in many low- and middle-income countries (per the World Bank definition; e.g. India, Brazil), which are unfortunately also seeing high rates of SARS-CoV-2 infection and COVID-induced mortality. This study motivates follow-up prospective research into the specific risk factors we have identified that appear to predispose some patients towards the after effects of COVID-19.

**Article summary – Strengths and limitations of this study:**

- This is the first study at a major healthcare center analyzing risk factors for post-viral clearance hospitalization of COVID-19 patients.
- This analysis uses augmented curation methods to identify complications and comorbidities from the physician notes, rather than relying upon ICD codes.
- The statistical analysis identifies specific comorbidities in the year preceding PCR diagnosis of SARS-CoV-2 which are associated with increased rates of post-viral clearance hospitalization.
- The dataset used for this study is limited to a single healthcare system, so the underlying clinical characteristics of the study population are biased to reflect the clinical characteristics of individuals that receive medical treatment in certain regions of the United States (Arizona, Florida, Minnesota).
- In this study, we use the first of two consecutive negative PCR tests to estimate the viral clearance date for each patient, however the true viral clearance date for each patient is unknown.

## Introduction

To date, 42.1 million people worldwide have been infected with SARS-CoV-2 with over 1 million deaths as of October 2020^1^. Although many patients survived the disease, there is increasing attention to the possibility of symptom development and hospitalization among patients who have previously “cleared” the virus, as indicated by 2+ consecutive negative PCR test results. Data on the long-term outcomes among cleared SARS-CoV-2 patients has been thus far limited and often poorly defined. Some patients labeled as cleared have been shown to have a new positive test after documented negative tests, a positive test after a period of quarantine, or develop new symptoms requiring hospitalization after documented negative tests^2–6^. The former case is most narrowly suggestive of delayed viral shedding, false positives, or re-infection while the latter case allows for a more broader interpretation of possible outcomes after clearance to also include delayed complications of the index hospitalization and manifestations of a systemic inflammatory response^7^. Here, we will refer to clearance as patients with a history of confirmed positive SARS-CoV-2 PCR and two subsequent consecutive negative PCR tests based on nasopharyngeal swabs.

A recent case series^8^ reported four patients with apparent SARS-CoV-2 reinfection after an index hospitalization, despite resolution of symptoms and radiographic abnormalities and two consecutive negative tests separated by a day. A single center cohort study of 414 patients with SARS-CoV-2 reported a reinfection rate of 16.7% among cleared patients^5^, while a second cohort study of 262 patients with SARS-CoV-2 reported a reinfection rate of 14.5% among cleared patients^4^. To date, details on the outcomes of patients who require hospitalization after documented clearance of SARS-CoV-2, however, remain unknown. These hospitalizations can have enormous implications beyond individual outcomes in shaping how we guide our public health strategy, allocate healthcare resources, and identify those at greatest risk.

The objective of this study is to characterize the cohort of COVID-19 patients who are admitted or readmitted to the hospital for post-clearance of the virus (**Figure 1**). Here, we define the clearance date of the virus as the first of 2+ consecutive negative PCR tests for SARS-CoV-2 infection. We compared this cohort of patients against COVID-19 patients who are hospitalized pre-clearance of the virus, but not post-clearance. We then used augmented curation to compare rates of COVID-19 associated complications in these two cohorts over the following time horizons: **(1) Pre-COVID:** Day-365 to Day-11 relative to the first positive PCR test for SARS-CoV-2, **(2) SARS-CoV-2 positive:** Day-10 relative to the first positive PCR test up to the estimated viral clearance date, and **(3) Post viral clearance:** Day 1 to Day 90 relative to the estimated viral clearance date. In particular, we considered the following complications: anemia, cardiac arrhythmias, pleural effusion, hypertension, acute respiratory distress syndrome/acute lung injury, heart failure, hyperglycemia, acute kidney injury, and respiratory failure. We ran statistical significance tests to identify complications which are enriched in the hospitalized post-clearance cohort across the three time periods. Finally, we explored the temporal distribution of phenotype occurrences in the clinical notes for both cohorts over the time window +/- 90 days relative to the clearance date of the virus.

**Figure 1:**
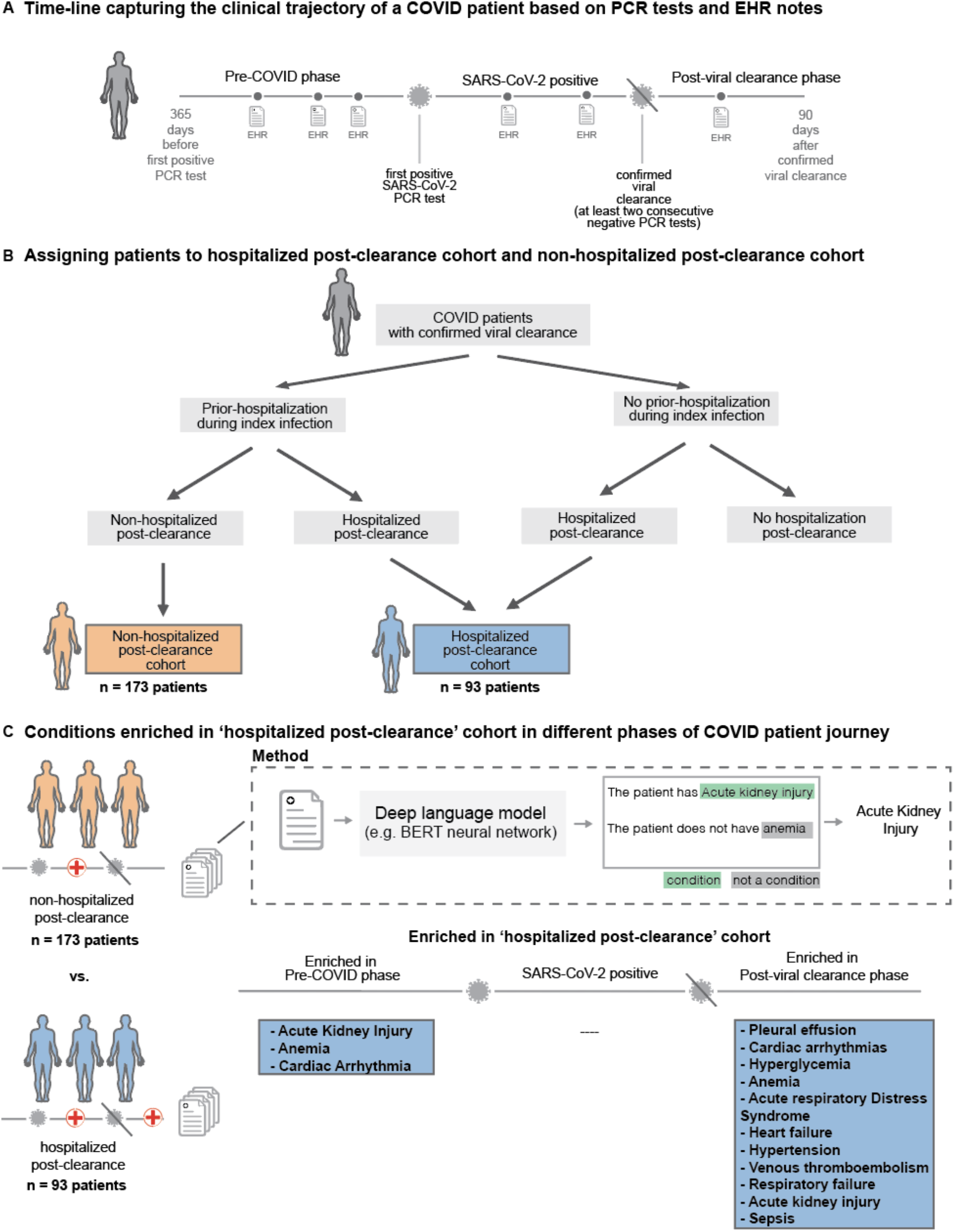
Overall study design. **(A)** Trajectory of a COVID-19 patient transitioning from pre-COVID (time up to 365 days before first positive PCR test) through the SARS-CoV-2 positive phase (interval after first positive test but before the first of two consecutive negative PCR test results) into the post viral clearance phase (period up to 90 days after the first of two negative PCR test results), **(B)** Demonstrates procedure for assigning patients to the hospitalized post-clearance cohort and non-hospitalized post-clearance cohort patients can be hospitalized at varying points in time including during the index infection (time from first positive PCR test results to first of two negative PCR test results) and following viral clearance - two cohorts are defined from the overall population, Hospitalized Post-Clearance Cohort in which patients are admitted or readmitted to the hospital following their estimated clearance date and Non-Hospitalized Post-Clearance Cohort in which patients are admitted during the index infection, but not following the estimated clearance date, **(C)** For each patient if the two defined cohorts a deep language (BERT) model is used to extracted phenotypes of interest from the clinical notes recorded between 365 days prior to infection and up to 90 days after clearance for each patient - occurrences of these phenotypes are stratified into pre-COVID, COVID pre-clearance, and COVID post-clearance time periods and statistical tests are run to find significant differences in phenotypes between the two cohorts.

## Methods

### Institutional Review Board (IRB)

This retrospective research was conducted under IRB 20–003278, ‘Study of COVID-19 patient characteristics with augmented curation of Electronic Health Records (EHR) to inform strategic and operational decisions’. For further information regarding the Mayo Clinic Institutional Review Board (IRB) policy, and its institutional commitment, membership requirements, review of research, informed consent, recruitment, vulnerable population protection, biologics, and confidentiality policy, please refer to www.mayo.edu/research/institutional-review-board/overview.

### Patient and Public Involvement

The development of the research question and outcome measures was informed by prior literature and information from the Centers for Disease Control and Prevention (CDC) on risk factors for severe COVID-19 illness^9^. No patients were involved in the design of the study, but physicians from the Mayo Clinic who are involved with the COVID research taskforce and the clinical care for COVID patients were involved with the study design and execution.

### Study design

Of 22,223 patients with at least one positive SARS-CoV-2 PCR test, 1,355 had two documented negative tests following their last positive test result with an estimated viral clearance date (date of the first negative test) more than 90 days prior to the date of this analysis (October 27, 2020). Among these 1,355 patients, 266 patients were admitted to the hospital, and 93 patients were admitted or readmitted to the hospital following the estimated viral clearance date. We define the ***hospitalized post-clearance*** cohort to consist of these 93 patients. We note that this cohort does not include patients who simply remained in the hospital following their estimated clearance date. On the other hand, 173 patients were admitted to the hospital pre-clearance, but were not admitted or readmitted to the hospital following the estimated clearance date. We refer to this group of 173 patients as the ***non-hospitalized post-clearance*** cohort. In **Figure 1**, we provide a schematic of the study design and the key findings from this study. In **Table 1**, we provide the general clinical characteristics for the two cohorts, including:

- **Demographics:** age, gender, race, ethnicity
- **Hospitalization status COVID Pre-clearance:** whether or not the patient was admitted to the hospital during the index COVID-19 infection,
- **Relative Clearance Date:** Number of days between first positive PCR test and first of two negative PCR tests.
- **Comorbidities:** Anemia, Asthma, Cancer, Chronic Kidney Disease (CKD), Hyperglycemia, Hypertension, Obesity, Obstructive sleep apnea, Type 1 diabetes mellitus (T1DM), Type 2 diabetes mellitus (T2DM).

**Table 1:**
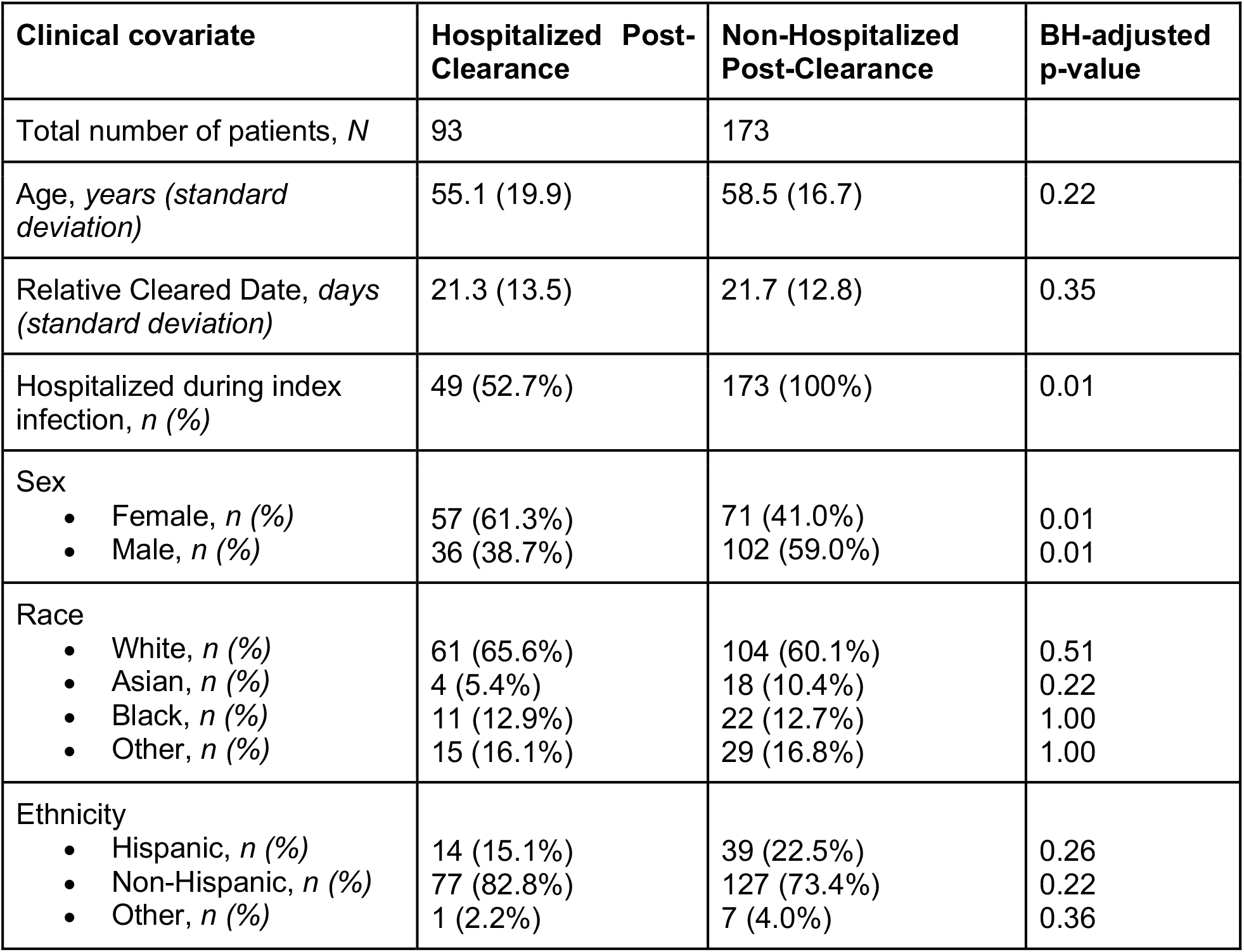
General characteristics of study population (patients who “cleared” after previously being SARS-CoV-2 positive) for hospitalized and non-hospitalized post-clearance cohorts. Demographic variables in the hospitalized and non-hospitalized post-clearance cohorts. The columns are: **(1) Hospitalized post-clearance:** clinical characteristics of the cohort of patients who are admitted or readmitted to the hospital following the estimated clearance date of SARS-CoV-2 infection, **(2) Non-Hospitalized post-clearance:** clinical characteristics of the control group of patients who are not admitted to the hospital following the estimated clearance date, **(3) BH-corrected P-value:** Benjamani-Hochberg corrected p-values using Fisher Exact test for comparisons of proportions and Mann-Whitney U test for continuous covariates.

| Clinical covariate | Hospitalized Post-Clearance | Non-Hospitalized Post-Clearance | BH-adjusted p-value |
| --- | --- | --- | --- |
| Total number of patients, *N* | 93 | 173 |  |
| Age, years (*standard deviation*) | 55.1 (19.9) | 58.5 (16.7) | 0.22 |
| Relative Cleared Date, days (*standard deviation*) | 21.3 (13.5) | 21.7 (12.8) | 0.35 |
| Hospitalized during index infection, *n* (%) | 49 (52.7%) | 173 (100%) | 0.01 |
| Sex<br>Female, *n* (%)<br>Male, *n* (%) | 57 (61.3%)<br>36 (38.7%) | 71 (41.0%)<br>102 (59.0%) | 0.01<br>0.01 |
| Race<br>White, *n* (%)<br>Asian, *n* (%)<br>Black, *n* (%)<br>Other, *n* (%) | 61 (65.6%)<br>4 (5.4%)<br>11 (12.9%)<br>15 (16.1%) | 104 (60.1%)<br>18 (10.4%)<br>22 (12.7%)<br>29 (16.8%) | 0.51<br>0.22<br>1.00<br>1.00 |
| Ethnicity<br>Hispanic, *n* (%)<br>Non-Hispanic, *n* (%)<br>Other, *n* (%) | 14 (15.1%)<br>77 (82.8%)<br>1 (2.2%) | 39 (22.5%)<br>127 (73.4%)<br>7 (4.0%) | 0.26<br>0.22<br>0.36 |

### Augmented curation of clinical notes

A BERT-based neural network^10^ was applied to identify phenotypes of interest in the clinical notes of the study population. The model was previously developed to classify the sentiment of general phenotypes^11^ and thrombotic event phenotypes^12^ in the encounter notes of COVID-19 patients. The categories of this classification model include: Yes (confirmed diagnosis), No (ruled out diagnosis), Maybe (possibility of disease), and Other (alternate context, e.g. family history of disease). This model was trained using nearly 250 different phenotypes and 18,500 sentences and achieves 93.6% overall accuracy and over 95% precision and recall for Yes/No sentiment classification^11^. For this study, the phenotypes of interest included all of the following comorbidities and complications along with their synonyms:

- **Comorbidities:** Anemia, Asthma, Cancer, Chronic Kidney Disease (CKD), Hyperglycemia, Hypertension, Obesity, Obstructive sleep apnea, Type 1 diabetes mellitus (T1DM), Type 2 diabetes mellitus (T2DM).
- **Complications:** Acute respiratory distress syndrome / acute lung injury (ARDS / ALI), Acute kidney injury (AKI), Cardiac arrest, Cardiac arrhythmias, Disseminated intravascular coagulation (DIC), Heart failure, Myocardial infarction, Pleural effusion, Pulmonary embolism, Respiratory failure, Sepsis, Septic shock, Stroke / Cerebrovascular incident, Venous thromboembolism / deep vein thrombosis (VTE / DVT).

We ran the model to classify the sentiment for all of the above phenotypes in all of the clinical notes for each patient in the time periods considered in this study. Only sentences containing a phenotype with a positive sentiment (labeled “Yes” by the model) with a confidence of 0.95 or above were deemed positive mentions. Repeated sentences for the same patient are ignored. For the SARS-CoV-2 positive phase and post-viral clearance phase, we only consider the presence or absence of a positive sentiment. For the pre-COVID time window, we consider thresholds of 1, 2, and 3 as the minimum number of positive mentions.

### SARS-CoV-2 PCR and IgG tests conducted by Mayo Clinic hospitals and health system

In the context of PCR testing, patients seen at Mayo Clinic in Rochester MN were tested by either a laboratory-developed test or the Roche cobas SARS-CoV-2 assay^13,14^. Patients seen at Mayo Clinic’s Florida hospitals and Arizona hospitals were tested using the Roche cobas test and Abbott diagnostic test respectively^15^. These SARS-CoV-2 PCR tests amplify different segments of the viral genome but are considered largely equivalent from the perspective of their analytical performance. The Logical Observation Identifiers Names and Codes (LOINC) code of the SARS-CoV-2 IgG test analyzed is 94563-4 (https://loinc.org/94563-4/)^16^. In the context of SARS-CoV-2 IgG test, patient serum samples were tested by Euroimmun Inc. Anti-SARS-CoV-2 IgG ELISA (Lubeck, Germany) according to the manufacturer instructions and as described previously on the Agility automated ELISA analyzer (Dynex Technologies, Inc., Chantilly, VA)^17^. This is a qualitative ELISA for detection of IgG-class antibodies against a recombinant version of the SARS-CoV-2 spike subunit 1 (S1) protein and has received Food and Drug Administration Emergency Use Authorization.

### Statistical significance tests

Fisher exact tests were applied pairwise across each of the covariates of interest in comparing each cohort to its respective control group, generating both a *p*-value and an Odds Ratio. This test was applied using the SciPy package^18^ in Python. *p*-values generated in this hypothesis were corrected using a Benjamini-Hochberg correction.

## Results

### COVID patients hospitalized post-viral clearance have higher rates of pre-COVID acute kidney injury, anemia, and cardiac arrhythmias as underlying conditions

In order to understand the patient characteristics that differentiated the ‘hospitalized post-clearance’ and ‘non-hospitalized post-clearance’ groups, we compared the clinical covariates in both the groups by analyzing phenotypes recorded with a positive sentiment in the patient history of both groups. In **Table 2**, we present the incidence of phenotypes which are enriched in the clinical notes during the pre-COVID time period, considering 1, 2, and 3 as the thresholds for distinct positive mentions. When the threshold is 1, we count all of the patients even with a single positive mention of the phenotype, and as the threshold increases, this criterion becomes more stringent and there is a monotonic decline.

**Table 2:**
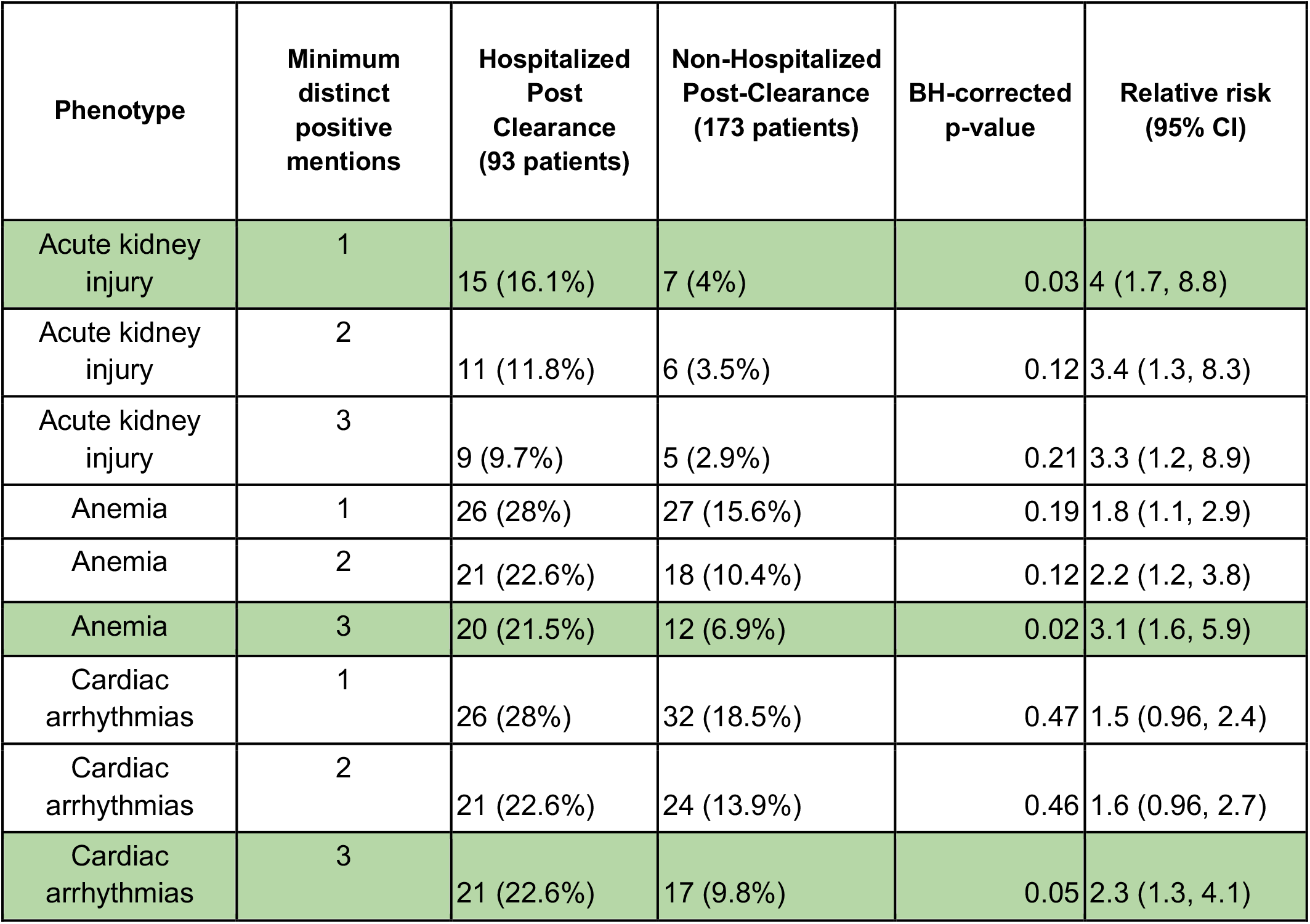
Enrichments of significant phenotypes in the Pre-COVID time horizon for 1, 2, and 3 clinical note thresholds (AKI, anemia, and cardiac arrhythmias). Phenotypes in the hospitalized and non-hospitalized post-clearance cohorts, along with results from statistical significance tests representing the time period one year prior up to 11 days prior to the first positive SARS-CoV-2 PCR test for a patient. Features which are significantly different between the two cohorts are highlighted in **green**. The columns are: **(1) Phenotype:** phenotype of interest, **(2) Minimum distinct positive mentions:** Minimum number of distinct mentions in the physician notes with a positive sentiment required to record a phenotype, **(3) Hospitalized post-clearance:** number of patients in the hospitalized post-clearance cohort with the phenotype recorded in the Pre-COVID time horizon, **(4) Non-Hospitalized post-clearance:** number of patients in the non-hospitalized post-clearance cohort with the phenotype recorded in the Pre-COVID time horizon, **(5) BH-corrected p-value:** Benjamani-Hochberg corrected p-values using Fisher Exact test for comparisons of proportions, **(6) Relative risk (95% CI):** Ratio of the phenotype incidence in the hospitalized post-clearance cohort to the phenotype incidence in the non-hospitalized post-clearance cohort, along with 95% confidence interval bounds.

We observe that with a threshold of 1 distinct positive mention, acute kidney injury (AKI) is enriched in the hospitalized post-clearance cohort (n: 15 (16.1%), relative risk: 4, 95% C.I.: [1.7, 8.8], p-value: 0.03). This suggests that patients with even a single positive mention of AKI may be at higher risk of poorer long-term prognosis, such as hospitalization post-clearance of the virus. In addition, with a threshold of 3 distinct positive mentions, the hospitalized post-clearance cohort is enriched for anemia (n: 20 (21.5%), relative risk: 3.1, 95% C.I.: [1.6, 5.9], p-value: 0.02) and cardiac arrhythmias (n: 21 (22.6%), relative risk: 2.3, 95% C.I.: [1.3, 4.1], p-value: 0.05). The fact that these phenotypes require more mentions to differentiate the two cohorts motivates the need to investigate whether these incidents are chronic or recurring, and how that may be linked with increased risk of post-clearance hospitalization.

In **Figure 2**, we present the temporal distribution of phenotypes in the clinical notes for acute kidney injury, anemia, and cardiac arrhythmias for the Pre-COVID time period. We observe that both cohorts have higher rates of reports for all three phenotypes in the time period of −50 days to −10 days relative to the first positive PCR testing date. In addition, we observe that acute kidney injury is reported more frequently in the clinical notes from −200 days to −280 days relative to the first positive PCR testing date. These clusters of notes may reflect groups of patients who were admitted to the hospital with these phenotypes around the same time. In **Supplementary Table S1**, we present the enrichment results for all of the pre-existing conditions that we considered, including those which are not significantly differentiated.

**Figure 2.**
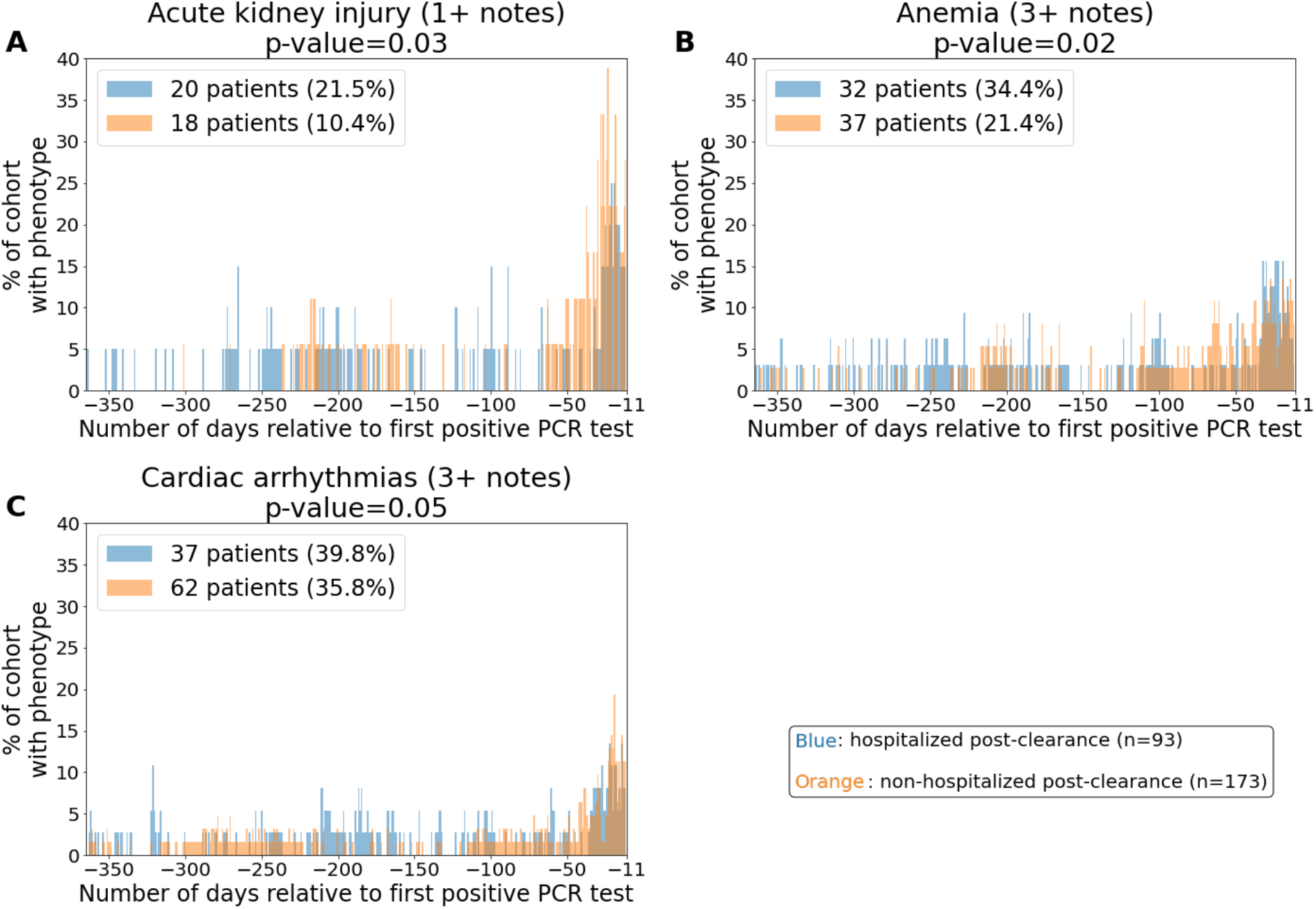
Distributions of comorbidities in the hospitalized and non-hospitalized cohorts for days-365 to -11 relative to the first positive PCR test. The phenotypes include: **(A)** Acute respiratory distress syndrome / acute lung injury (ARDS / ALI), **(B)** Anemia, and **(C)** Cardiac arrhythmias. In the subplot for a single phenotype, the x-axis corresponds to the date relative to the first positive PCR date, and the y-axis corresponds to the percentage of patients with mentions of the phenotype with positive sentiment in their clinical notes on that relative date. In the title for each plot, we show the (phenotype, threshold for positive sentiment mentions) pair which is enriched in the pre-COVID phase, along with the associated p-value. In the legend for each plot, we show the number and percentage of patients with the phenotype for the hospitalized post-clearance cohort in **blue** and for the non-hospitalized post-clearance cohort in **orange**.

### Post-clearance of SARS-CoV-2 virus, COVID patients experience complications including cardiovascular, renal, pulmonary, and immunological conditions

In **Table 3**, we present the rates of complications for the two cohorts of interest for the two time periods following PCR diagnosis of SARS-CoV-2: (1) SARS-CoV-2 positive phase, and (2) post-viral clearance phase. For the post-viral clearance phase, we observe the following complications at higher rates in the hospitalized post-clearance cohort: acute kidney injury, anemia, acute respiratory distress syndrome / acute lung injury, cardiac arrhythmias, heart failure, hyperglycemia, hypertension, pleural effusion, respiratory failure, sepsis, and venous thromboembolism / deep vein thrombosis. Among these, the most significantly enriched phenotypes are: pleural effusion (relative risk: 4.2, 95% C.I.: [2.1, 7.9], p-value: 1.2E-4), cardiac arrhythmias (relative risk: 5.3, 95% C.I.: [2.1, 12], p-value: 9.3E-4), and hyperglycemia (relative risk: Infinity, 95% C.I.: [1.8, 5,400], p-value: 1.0E-3).

**Table 3.**
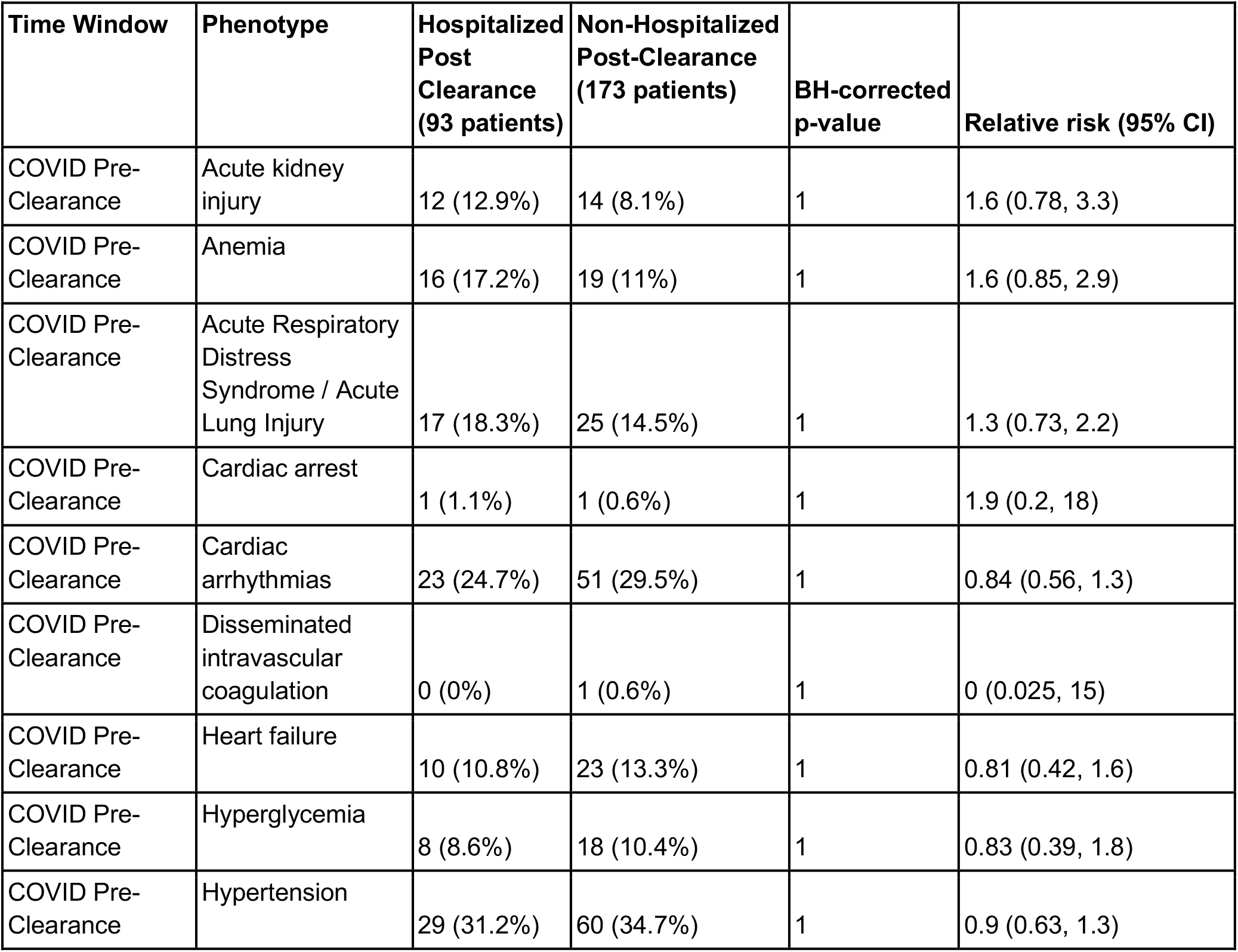
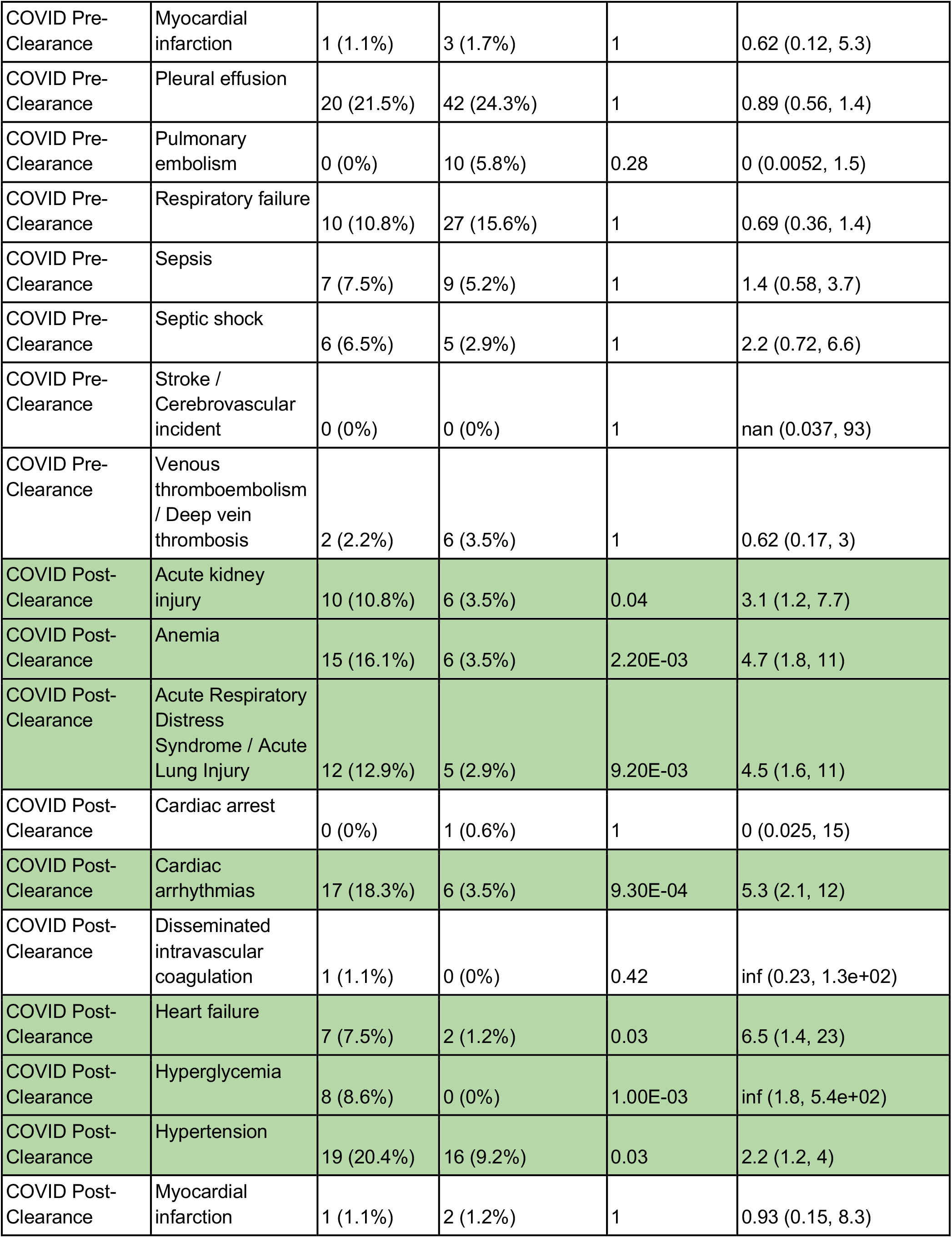
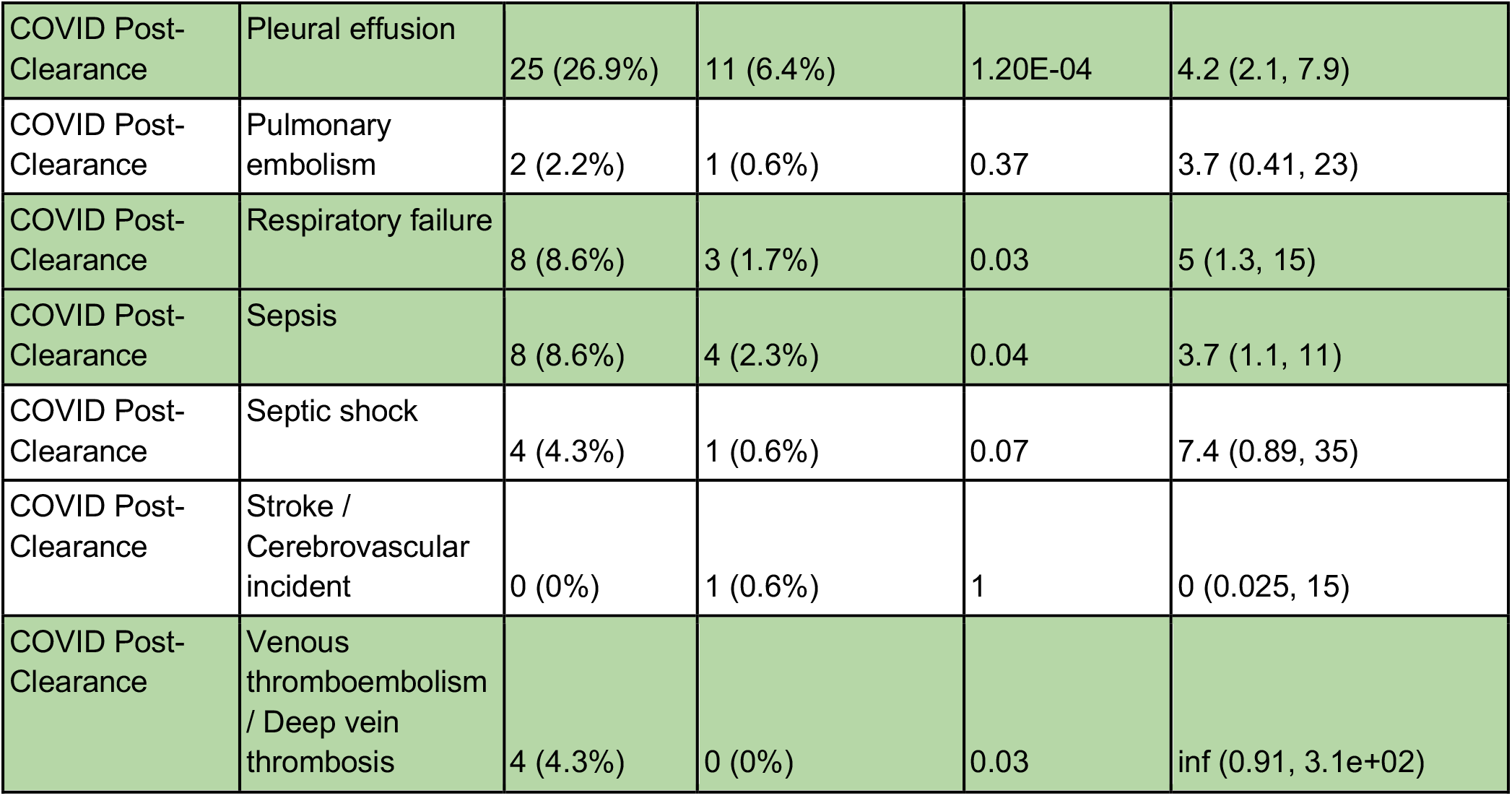
Comparison of complications in the hospitalized and non-hospitalized post-clearance cohorts. Complications in the hospitalized and non-hospitalized post-clearance cohorts, along with results from statistical significance tests stratified by time periods including: pre-covid infection, between SARS-CoV-2 infection and clearance, and post-COVID infection. Phenotypes are identified in the physician notes via the neural network model with a minimum number of 1+ distinct mentions with a positive sentiment in the time horizon required to record a phenotype. Features which are significantly different between the two cohorts are highlighted in **green**. The columns are: **(1) Time window:** time window of interest, **(2) Phenotype:** phenotype of interest, **(3) Hospitalized post-clearance:** number of patients in the hospitalized post-clearance cohort with the phenotype recorded in the specified time horizon, **(4) Non-Hospitalized post-clearance:** number of patients in the non-hospitalized post-clearance cohort with the phenotype recorded in the specified time horizon, **(5) BH-corrected p-value:** Benjamani-Hochberg corrected p-values using Fisher Exact test for comparisons of proportions, **(6) Relative risk (95% CI):** Ratio of the phenotype incidence in the hospitalized post-clearance cohort to the phenotype incidence in the non-hospitalized post-clearance cohort, along with 95% confidence interval bounds.

| Time Window | Phenotype | Hospitalized Post Clearance (93 patients) | Non-Hospitalized Post-Clearance (173 patients) | BH-corrected p-value | Relative risk (95% CI) |
| --- | --- | --- | --- | --- | --- |
| COVID Pre-Clearance | Acute kidney injury | 12 (12.9%) | 14 (8.1%) | 1 | 1.6 (0.78, 3.3) |
| COVID Pre-Clearance | Anemia | 16 (17.2%) | 19 (11%) | 1 | 1.6 (0.85, 2.9) |
| COVID Pre-Clearance | Acute Respiratory Distress Syndrome / Acute Lung Injury | 17 (18.3%) | 25 (14.5%) | 1 | 1.3 (0.73, 2.2) |
| COVID Pre-Clearance | Cardiac arrest | 1 (1.1%) | 1 (0.6%) | 1 | 1.9 (0.2, 18) |
| COVID Pre-Clearance | Cardiac arrhythmias | 23 (24.7%) | 51 (29.5%) | 1 | 0.84 (0.56, 1.3) |
| COVID Pre-Clearance | Disseminated intravascular coagulation | 0 (0%) | 1 (0.6%) | 1 | 0 (0.025, 15) |
| COVID Pre-Clearance | Heart failure | 10 (10.8%) | 23 (13.3%) | 1 | 0.81 (0.42, 1.6) |
| COVID Pre-Clearance | Hyperglycemia | 8 (8.6%) | 18 (10.4%) | 1 | 0.83 (0.39, 1.8) |
| COVID Pre-Clearance | Hypertension | 29 (31.2%) | 60 (34.7%) | 1 | 0.9 (0.63, 1.3) |
| COVID Pre-Clearance | Myocardial infarction | 1 (1.1%) | 3 (1.7%) | 1 | 0.62 (0.12, 5.3) |
| COVID Pre-Clearance | Pleural effusion | 20 (21.5%) | 42 (24.3%) | 1 | 0.89 (0.56, 1.4) |
| COVID Pre-Clearance | Pulmonary embolism | 0 (0%) | 10 (5.8%) | 0.28 | 0 (0.0052, 1.5) |
| COVID Pre-Clearance | Respiratory failure | 10 (10.8%) | 27 (15.6%) | 1 | 0.69 (0.36, 1.4) |
| COVID Pre-Clearance | Sepsis | 7 (7.5%) | 9 (5.2%) | 1 | 1.4 (0.58, 3.7) |
| COVID Pre-Clearance | Septic shock | 6 (6.5%) | 5 (2.9%) | 1 | 2.2 (0.72, 6.6) |
| COVID Pre-Clearance | Stroke / Cerebrovascular incident | 0 (0%) | 0 (0%) | 1 | nan (0.037, 93) |
| COVID Pre-Clearance | Venous thromboembolism / Deep vein thrombosis | 2 (2.2%) | 6 (3.5%) | 1 | 0.62 (0.17, 3) |
| COVID Post-Clearance | Acute kidney injury | 10 (10.8%) | 6 (3.5%) | 0.04 | 3.1 (1.2, 7.7) |
| COVID Post-Clearance | Anemia | 15 (16.1%) | 6 (3.5%) | 2.20E-03 | 4.7 (1.8, 11) |
| COVID Post-Clearance | Acute Respiratory Distress Syndrome / Acute Lung Injury | 12 (12.9%) | 5 (2.9%) | 9.20E-03 | 4.5 (1.6, 11) |
| COVID Post-Clearance | Cardiac arrest | 0 (0%) | 1 (0.6%) | 1 | 0 (0.025, 15) |
| COVID Post-Clearance | Cardiac arrhythmias | 17 (18.3%) | 6 (3.5%) | 9.30E-04 | 5.3 (2.1, 12) |
| COVID Post-Clearance | Disseminated intravascular coagulation | 1 (1.1%) | 0 (0%) | 0.42 | inf (0.23, 1.3e+02) |
| COVID Post-Clearance | Heart failure | 7 (7.5%) | 2 (1.2%) | 0.03 | 6.5 (1.4, 23) |
| COVID Post-Clearance | Hyperglycemia | 8 (8.6%) | 0 (0%) | 1.00E-03 | inf (1.8, 5.4e+02) |
| COVID Post-Clearance | Hypertension | 19 (20.4%) | 16 (9.2%) | 0.03 | 2.2 (1.2, 4) |
| COVID Post-Clearance | Myocardial infarction | 1 (1.1%) | 2 (1.2%) | 1 | 0.93 (0.15, 8.3) |
| COVID Post-Clearance | Pleural effusion | 25 (26.9%) | 11 (6.4%) | 1.20E-04 | 4.2 (2.1, 7.9) |
| COVID Post-Clearance | Pulmonary embolism | 2 (2.2%) | 1 (0.6%) | 0.37 | 3.7 (0.41, 23) |
| COVID Post-Clearance | Respiratory failure | 8 (8.6%) | 3 (1.7%) | 0.03 | 5 (1.3, 15) |
| COVID Post-Clearance | Sepsis | 8 (8.6%) | 4 (2.3%) | 0.04 | 3.7 (1.1, 11) |
| COVID Post-Clearance | Septic shock | 4 (4.3%) | 1 (0.6%) | 0.07 | 7.4 (0.89, 35) |
| COVID Post-Clearance | Stroke / Cerebrovascular incident | 0 (0%) | 1 (0.6%) | 1 | 0 (0.025, 15) |
| COVID Post-Clearance | Venous thromboembolism / Deep vein thrombosis | 4 (4.3%) | 0 (0%) | 0.03 | inf (0.91, 3.1e+02) |

In **Figure 3**, we show rates of these phenotypes over the +/- 90 time window centered around the estimated viral clearance date for each patient. For most of these complications, we observe for the case cohort that the majority of reports in the clinical notes occur within the first three weeks following the estimated viral clearance date. An exception is for pleural effusions, which occurs relatively frequently in the notes of the case cohort in the two months following the clearance date. In addition, we observe moderate amounts of reports for cardiac arrhythmias, hyperglycemia, anemia, acute respiratory distress syndrome / acute lung injury, hypertension, and venous thromboembolism / deep vein thrombosis around 50 to 70 days post-clearance.

**Figure 3.**
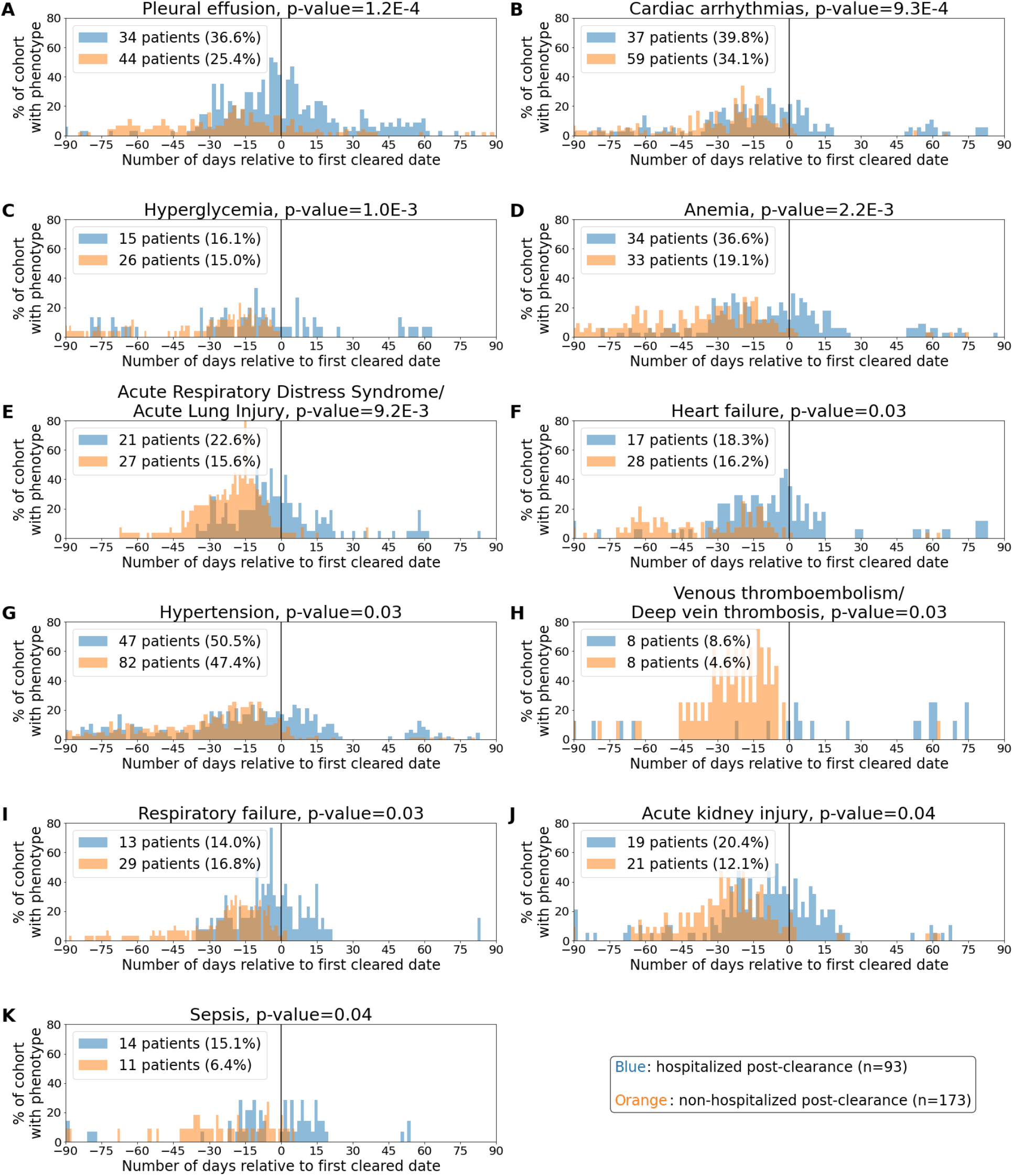
Distributions of complications in the hospitalized and non-hospitalized cohorts for days-90 to 90 relative to the virus clearance date. The phenotypes include: **(A)** Pleural effusion, **(B)** Cardiac arrhythmias, **(C)** Hyperglycemia, **(D)** Anemia, **(E)** Acute respiratory distress syndrome / acute lung injury (ARDS / ALI), **(F)** Heart failure, **(G)** Hypertension, **(H)** Venous thromboembolism / Deep Vein Thrombosis, **(I)** Respiratory Failure, **(J)** Acute Kidney Injury, and **(K)** Sepsis. In the subplot for a single phenotype, the x-axis corresponds to the date relative to the virus clearance date, and the y-axis corresponds to the percentage of patients with mentions of the phenotype with positive sentiment in their clinical notes on that relative date. The clearance date (day = 0) is indicated by a vertical line. In the title for each plot, we show the phenotype which is enriched in the post-clearance phase, along with the associated p-value. In the legend for each plot, we show the number and percentage of patients with the phenotype from days -90 to 90 (relative to clearance date) for the hospitalized post-clearance cohort in **blue** and for the non-hospitalized post-clearance cohort in **orange**.

## Discussion

The topic of hospitalization and readmission in COVID-19 patients has been under major focus for mitigating national healthcare costs, particularly among 30-day readmissions that account for $17 billion in avoidable Medicare expenditures^19^. Prior studies have investigated hospitalization and readmission after recovery from other acute illnesses, such as influenza and community-acquired pneumonia, heart failure, and decompensated cirrhosis. In the case of influenza, the most common cause of short-term re-admission is due to non-influenza pneumonia, thought to result from complex interactions between the recovering host immune system and inflammatory response, reminiscent to a limited extent of the systemic inflammatory response seen in SARS-CoV-2. These outcomes are associated with notably higher costs of care and increased mortality^20^. Given the widespread prevalence of SARS-CoV-2, similar outcomes warrant detailed studies in patients hospitalized after SARS-CoV-2 clearance.

In this study, we found that patients hospitalized after SARS-CoV-2 clearance had significantly higher odds of having comorbidities of anemia as well as experiencing serious complications of ARDS/ALI, AKI, pleural effusion, and sepsis compared to hospitalized controls. Approximately a quarter (24.5%) of these patients required ICU admission. In addition, we found that hypertension, asthma, obstructive sleep apnea, obesity, and diabetes were not increased among patients hospitalized after clearance compared to controls. These are co-morbidities that have been previously well-described to be associated with poorer outcomes during an index SARS-CoV-2 infection^21^. One possible hypothesis for these findings is that many of these hospitalizations post-clearance were not due to re-infection per se, but rather secondary to a systemic inflammatory response, resulting in the observed increased frequency of ARDS and sepsis noted in the cases, for which patients with a higher frailty index may be more susceptible. Patients hospitalized after clearance had increased odds of having anemia compared to their respective controls. It has been well-studied that patients with anemia, independent of other health conditions, are at higher risk of frailty and mortality, particularly in older patients^22^. Additionally, a limitation of our study is that our algorithm did not include all forms of obstructive airway disease, including COPD, which has been indicated to increase mortality in COVID-19^23^. Despite these caveats, our finding that pre-COVID anemia is endemic or highly prevalent in several low- and middle-income countries^23^, where COVID-19 is unfortunately a raging pandemic (e.g. India and Brazil) is especially concerning from the standpoint of higher post-viral clearance hospitalization risk in COVID patients.

A point of interest in explaining heterogeneous outcomes as regards hospital admission following SARS-CoV-2 viral clearance is the development of IgG antibodies post-clearance. In the context of this study 16 of 23 patients (69.5%) in the *hospitalized post-clearance cohort* tested for IgG antibodies test positive; this is compared to a rate of 19 of 23 patients (82.6%) in the *non-hospitalized post-clearance cohort*. The difference between these groups (16/23 vs. 19/23; fisher exact p-value 0.49) is not significant in this case. However, as the antibody testing data increases further investigation into the relationship between IgG seropositivity and complications following SARS-CoV-2 clearance may be warranted.

There are several limitations of this study due to the observational nature of the data. First, we are not controlling for PCR testing frequency, so the study population which receives access to multiple PCR tests to confirm SARS-CoV-2 infection clearance may not be representative of COVID-19 patient population in general. In addition, the phenotypes of complications and comorbidities which are identified in the notes with positive sentiment may In addition, the control groups for the hospitalized post-clearance cohorts were not propensity matched due to limited cohort size. However, patients in the hospitalized cohort did not differ in demographic characteristics, and compared to prior investigations on this subject, the size of our cohort is a strength of the study. Next, the current time points chosen to align the patient journeys are anchored by the first positive PCR test and the first of the two or more negative tests. Aligned the patient journeys based on their hospitalization events can help identify the conditions that are enriched with each hospitalization event. Finally, there is a limitation of the augmented curation model which is used to identify complications and comorbidities, because it only determines whether or not the phenotype was mentioned with positive sentiment in a clinical note, and does not take into account the temporal component. Therefore, although we know that certain phenotypes are enriched in the cohorts which are admitted to the hospital/ICU post-clearance, the dates of these complications are uncertain. There is research ongoing to develop natural language processing (NLP) based neural network models which can differentiate effectively between active and historical phenotype diagnoses, which could strengthen these conclusions for future analyses.

The phenotypes described here relate to EHR based mentions of conditions. Future work would include a complementary analysis based on lab tests relevant to the identified phenotypes such as estimated Glomerular Filtration Rate, Blood Urea Nitrogen test, Hemoglobin, Hematocrit, Alanine transaminase (ALT), Aspartate transaminase (AST) and Bilirubin. A longitudinal analysis integrating clinical notes-based augmented curation and lab-measurements will be useful in formulating a case definition of long-term COVID-19 (long-COVID). Overall, our finding of the long-term adverse effects of COVID-19 motivates the need to understand the biological and mechanistic underpinning of the SARS-CoV-2 driven long-term adverse effects in order to find appropriate prophylactic and therapeutic interventions. Finally, this study also emphasizes the need for detailed curation of structured and unstructured clinical data to better understand the dynamics of viral clearance, underlying conditions, and long-term complications.

## Data Availability

After publication, the data will be made available to others upon reasonable requests to the corresponding author. A proposal with detailed description of study objectives and statistical analysis plan will be needed for evaluation of the reasonability of requests. Deidentified data will be provided after approval from the corresponding author and the Mayo Clinic standard IRB process for such requests.

## Author Contributions

CP and AV contributed to the study design, project supervision, and writing. ER and CK performed the formal analysis and contributed to the writing. AP, NK, and GB contributed to the statistical analysis and validated the augmented curation models. AA and RB conceptualized and trained the augmented models, and RB contributed to the writing (review and editing). JO and ADB contributed to the investigation and writing (review and editing). VS conceived the study design, contributed to the investigation, project administration, project supervision, and writing (review and editing).

## Data Availability

After publication, the data will be made available to others upon reasonable requests to the corresponding author. A proposal with detailed description of study objectives and statistical analysis plan will be needed for evaluation of the reasonability of requests. Deidentified data will be provided after approval from the corresponding author and the Mayo Clinic’s standard IRB process for such requests.

## Supplementary Material

**Table S1.**
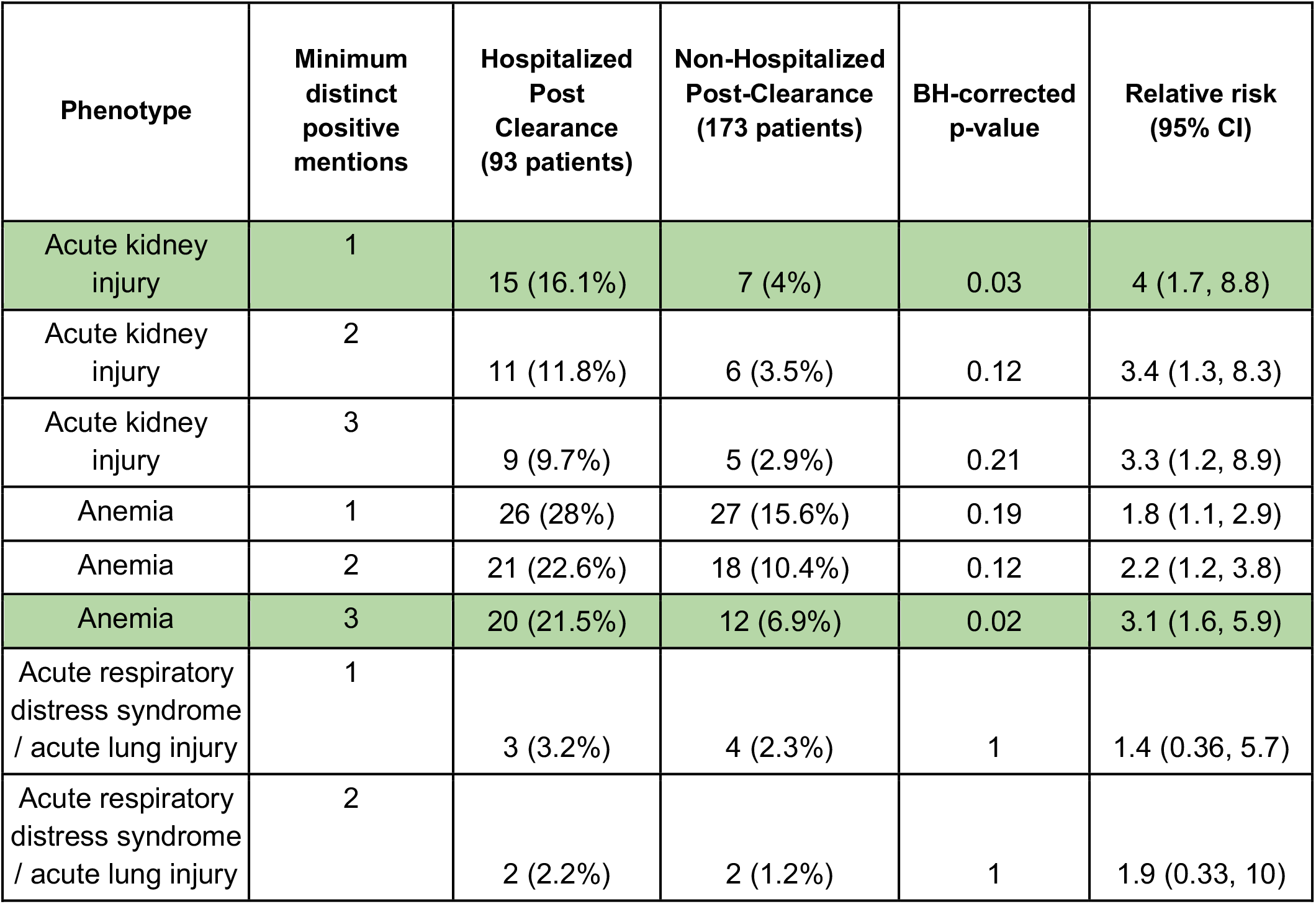
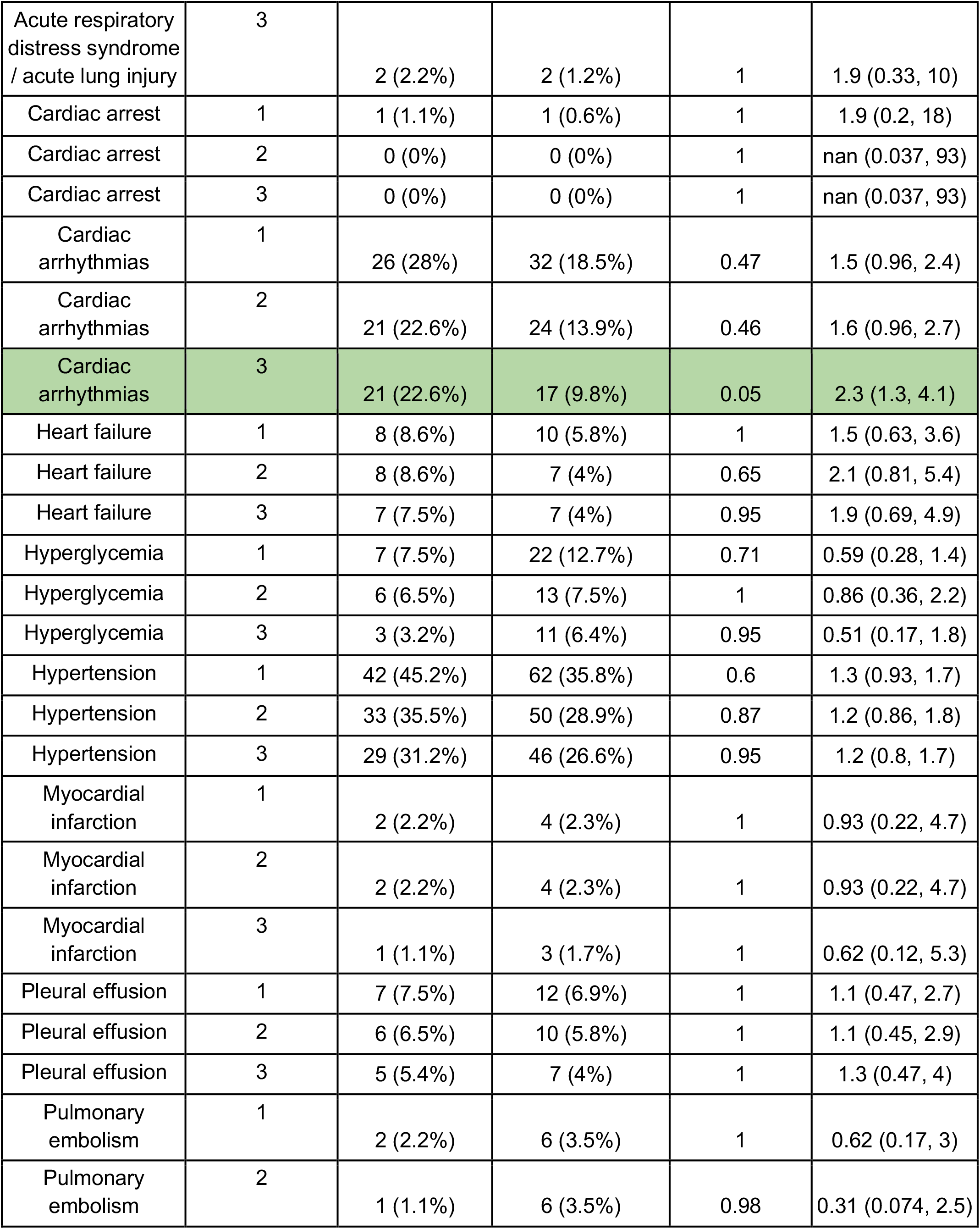
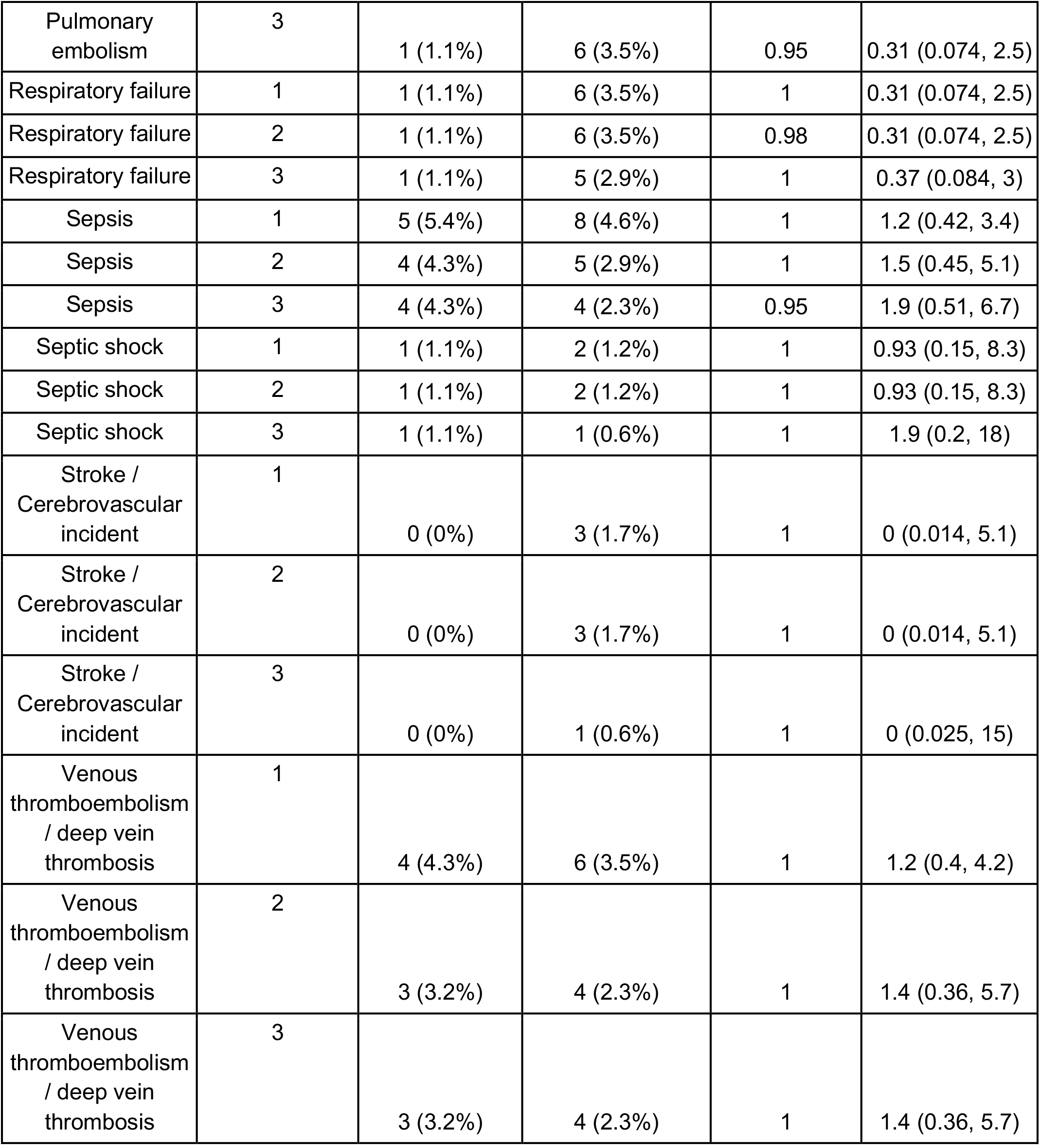
Enrichments of all phenotypes in the Pre-COVID time horizon for 1, 2, and 3 note thresholds. Phenotypes in the hospitalized and non-hospitalized post-clearance cohorts, along with results from statistical significance tests the time period one year prior to up to 11 days prior to the first positive SARS-CoV-2 PCR test for a patient. Features which are significantly different between the two cohorts are highlighted in **green**. The columns are: **(1) Phenotype:** phenotype of interest, **(2) Minimum distinct positive mentions:** Minimum number of distinct mentions in the physician notes with a positive sentiment required to record a phenotype, **(3) Hospitalized post-clearance:** number of patients in the hospitalized post-clearance cohort with the phenotype recorded in the Pre-COVID time horizon, **(4) Non-Hospitalized post-clearance:** number of patients in the non-hospitalized post-clearance cohort with the phenotype recorded in the Pre-COVID time horizon, **(5) BH-corrected p-value:** Benjamani-Hochberg corrected p-values using Fisher Exact test for comparisons of proportions, **(6) Relative risk (95% CI):** Ratio of the phenotype incidence in the hospitalized post-clearance cohort to the phenotype incidence in the non-hospitalized post-clearance cohort, along with 95% confidence interval bounds.

