## Supplementary Material for "Pre-existing conditions are associated with COVID patients’ hospitalization, despite confirmed clearance of SARS-CoV-2 virus"

**Table S1. Enrichments of all phenotypes in the Pre-COVID time horizon for 1, 2, and 3 note thresholds.** Phenotypes in the hospitalized and non-hospitalized post-clearance cohorts, along with results from statistical significance tests the time period one year prior to up to 11 days prior to the first positive SARS-CoV-2 PCR test for a patient. Features which are significantly different between the two cohorts are highlighted in green. The columns are: **(1) Phenotype:** phenotype of interest, **(2) Minimum distinct positive mentions:** Minimum number of distinct mentions in the physician notes with a positive sentiment required to record a phenotype, **(3) Hospitalized post-clearance:** number of patients in the hospitalized post-clearance cohort with the phenotype recorded in the Pre-COVID time horizon, **(4) Non-Hospitalized post-clearance:** number of patients in the non-hospitalized post-clearance cohort with the phenotype recorded in the Pre-COVID time horizon, **(5) BH-corrected p-value:** Benjamani-Hochberg corrected p-values using Fisher Exact test for comparisons of proportions, **(6) Relative risk (95% CI):** Ratio of the phenotype incidence in the hospitalized post-clearance cohort to the phenotype incidence in the non-hospitalized post-clearance cohort, along with 95% confidence interval bounds.

| Phenotype | Minimum distinct positive mentions | Hospitalized Post Clearance (93 patients) | Non-Hospitalized Post-Clearance (173 patients) | BH-corrected p-value | Relative risk (95% CI) |
|---|---|---|---|---|---|
| Acute kidney injury | 1 | 15 (16.1%) | 7 (4%) | 0.03 | 4 (1.7, 8.8) |
| Acute kidney injury | 2 | 11 (11.8%) | 6 (3.5%) | 0.12 | 3.4 (1.3, 8.3) |
| Acute kidney injury | 3 | 9 (9.7%) | 5 (2.9%) | 0.21 | 3.3 (1.2, 8.9) |
| Anemia | 1 | 26 (28%) | 27 (15.6%) | 0.19 | 1.8 (1.1, 2.9) |
| Anemia | 2 | 21 (22.6%) | 18 (10.4%) | 0.12 | 2.2 (1.2, 3.8) |
| Anemia | 3 | 20 (21.5%) | 12 (6.9%) | 0.02 | 3.1 (1.6, 5.9) |
| Acute respiratory distress syndrome / acute lung injury | 1 | 3 (3.2%) | 4 (2.3%) | 1 | 1.4 (0.36, 5.7) |
| Acute respiratory distress syndrome / acute lung injury | 2 | 2 (2.2%) | 2 (1.2%) | 1 | 1.9 (0.33, 10) |
| Acute respiratory distress syndrome / acute lung injury | 3 | 2 (2.2%) | 2 (1.2%) | 1 | 1.9 (0.33, 10) |
| Cardiac arrest | 1 | 1 (1.1%) | 1 (0.6%) | 1 | 1.9 (0.2, 18) |

| | | | | | |
|---|---|---|---|---|---|
| Cardiac arrest | 2 | 0 (0%) | 0 (0%) | 1 | nan (0.037, 93) |
| Cardiac arrest | 3 | 0 (0%) | 0 (0%) | 1 | nan (0.037, 93) |
| Cardiac arrhythmias | 1 | 26 (28%) | 32 (18.5%) | 0.47 | 1.5 (0.96, 2.4) |
| Cardiac arrhythmias | 2 | 21 (22.6%) | 24 (13.9%) | 0.46 | 1.6 (0.96, 2.7) |
| Cardiac arrhythmias | 3 | 21 (22.6%) | 17 (9.8%) | 0.05 | 2.3 (1.3, 4.1) |
| Heart failure | 1 | 8 (8.6%) | 10 (5.8%) | 1 | 1.5 (0.63, 3.6) |
| Heart failure | 2 | 8 (8.6%) | 7 (4%) | 0.65 | 2.1 (0.81, 5.4) |
| Heart failure | 3 | 7 (7.5%) | 7 (4%) | 0.95 | 1.9 (0.69, 4.9) |
| Hyperglycemia | 1 | 7 (7.5%) | 22 (12.7%) | 0.71 | 0.59 (0.28, 1.4) |
| Hyperglycemia | 2 | 6 (6.5%) | 13 (7.5%) | 1 | 0.86 (0.36, 2.2) |
| Hyperglycemia | 3 | 3 (3.2%) | 11 (6.4%) | 0.95 | 0.51 (0.17, 1.8) |
| Hypertension | 1 | 42 (45.2%) | 62 (35.8%) | 0.6 | 1.3 (0.93, 1.7) |
| Hypertension | 2 | 33 (35.5%) | 50 (28.9%) | 0.87 | 1.2 (0.86, 1.8) |
| Hypertension | 3 | 29 (31.2%) | 46 (26.6%) | 0.95 | 1.2 (0.8, 1.7) |
| Myocardial infarction | 1 | 2 (2.2%) | 4 (2.3%) | 1 | 0.93 (0.22, 4.7) |
| Myocardial infarction | 2 | 2 (2.2%) | 4 (2.3%) | 1 | 0.93 (0.22, 4.7) |
| Myocardial infarction | 3 | 1 (1.1%) | 3 (1.7%) | 1 | 0.62 (0.12, 5.3) |
| Pleural effusion | 1 | 7 (7.5%) | 12 (6.9%) | 1 | 1.1 (0.47, 2.7) |
| Pleural effusion | 2 | 6 (6.5%) | 10 (5.8%) | 1 | 1.1 (0.45, 2.9) |
| Pleural effusion | 3 | 5 (5.4%) | 7 (4%) | 1 | 1.3 (0.47, 4) |
| Pulmonary embolism | 1 | 2 (2.2%) | 6 (3.5%) | 1 | 0.62 (0.17, 3) |
| Pulmonary embolism | 2 | 1 (1.1%) | 6 (3.5%) | 0.98 | 0.31 (0.074, 2.5) |
| Pulmonary embolism | 3 | 1 (1.1%) | 6 (3.5%) | 0.95 | 0.31 (0.074, 2.5) |
| Respiratory failure | 1 | 1 (1.1%) | 6 (3.5%) | 1 | 0.31 (0.074, 2.5) |
| Respiratory failure | 2 | 1 (1.1%) | 6 (3.5%) | 0.98 | 0.31 (0.074, 2.5) |

| | | | | | |
|---|---|---|---|---|---|
| Respiratory failure | 3 | 1 (1.1%) | 5 (2.9%) | 1 | 0.37 (0.084, 3) |
| Sepsis | 1 | 5 (5.4%) | 8 (4.6%) | 1 | 1.2 (0.42, 3.4) |
| Sepsis | 2 | 4 (4.3%) | 5 (2.9%) | 1 | 1.5 (0.45, 5.1) |
| Sepsis | 3 | 4 (4.3%) | 4 (2.3%) | 0.95 | 1.9 (0.51, 6.7) |
| Septic shock | 1 | 1 (1.1%) | 2 (1.2%) | 1 | 0.93 (0.15, 8.3) |
| Septic shock | 2 | 1 (1.1%) | 2 (1.2%) | 1 | 0.93 (0.15, 8.3) |
| Septic shock | 3 | 1 (1.1%) | 1 (0.6%) | 1 | 1.9 (0.2, 18) |
| Stroke / Cerebrovascular incident | 1 | 0 (0%) | 3 (1.7%) | 1 | 0 (0.014, 5.1) |
| Stroke / Cerebrovascular incident | 2 | 0 (0%) | 3 (1.7%) | 1 | 0 (0.014, 5.1) |
| Stroke / Cerebrovascular incident | 3 | 0 (0%) | 1 (0.6%) | 1 | 0 (0.025, 15) |
| Venous thromboembolism / deep vein thrombosis | 1 | 4 (4.3%) | 6 (3.5%) | 1 | 1.2 (0.4, 4.2) |
| Venous thromboembolism / deep vein thrombosis | 2 | 3 (3.2%) | 4 (2.3%) | 1 | 1.4 (0.36, 5.7) |
| Venous thromboembolism / deep vein thrombosis | 3 | 3 (3.2%) | 4 (2.3%) | 1 | 1.4 (0.36, 5.7) |
